# Recurrent *Plasmodium falciparum* parasitemia and drug resistance mutations during intermittent preventive treatment of malaria in pregnancy in Uganda

**DOI:** 10.1101/2025.10.18.25338277

**Authors:** Jimmy Kizza, Thomas Katairo, Abel Kakuru, Bienvenu Nsengimaana, Trevor Esilu, Innocent Wiringilimaana, Francis D Semakuba, Inna Gerlovina, Nicholas Hathaway, Jessica Briggs, Stephen Tukwasibwe, Steven M. Kiwuwa, Moses R. Kamya, Joaniter I. Nankabirwa, Grant Dorsey, Philip J. Rosenthal

**Affiliations:** Infectious Diseases Research Collaboration, Kampala, Uganda; Busitema University, Tororo, Uganda; University of California, San Francisco, CA, USA; Makerere University, Kampala, Uganda

**Keywords:** Malaria, *Plasmodium falciparum*, intermittent preventive treatment, pregnancy, sulfadoxine-pyrimethamine, dihydroartemisinin-piperaquine, resistance, Uganda

## Abstract

**Background:** Intermittent preventive treatment with monthly sulfadoxine-pyrimethamine (IPTp-SP) is recommended during pregnancy in malaria-endemic countries. However, widespread resistance of *Plasmodium falciparum* to SP has compromised its efficacy, and the alternative dihydroartemisinin-piperaquine (DP) is under study. Potential selection of drug resistance is of interest.

**Methods:** We sequenced 1377 samples collected from pregnant women enrolled in a trial comparing monthly SP, DP, and DP+SP for IPTp in Busia, Uganda and with asymptomatic parasitemia at the time of IPTp administration. We characterized known markers of drug resistance and assessed the 28-day cumulative risk of recurrent parasitemia, with genotyping to distinguish recrudescence from new infections.

**Results:** Among 771 samples collected on the day IPTp was initiated, the prevalences of five resistance mutations in *P. falciparum* dihydrofolate reductase (PfDHFR) and dihydropteroate synthase (PfDHPS) were nearly 100%, and the PfDHFR I164L and PfDHPS A581G mutations, associated with high-level resistance, had combined prevalence of 26.5%. The cumulative risks of recurrent parasitemia (SP 57.8%, DP 4.1%, DP+SP 3.9%), symptomatic malaria (SP 9.3%, DP 1.1%, DP+SP 0.3%), and recrudescent parasitemia (SP 40.1%, DP 2.0%, DP+SP 0.8%) were all significantly greater in the SP arm, with risks greatest in primigravidae. In the IPT-SP arm, the combined prevalence of the PfDHFR I164L and PfDHPS A581G mutations increased significantly from 24.9% at initiation of IPTp to 35.2% after receipt of IPTp-SP. Infection with mutant parasites was associated with non-significant increases in risks of recrudescence.

**Conclusions:** IPTp-SP had poor preventive efficacy and selected for increased drug resistance, questioning the value of this intervention.

## INTRODUCTION

Malaria, in particular *Plasmodium falciparum* infection, remains a major health burden in sub-Saharan Africa [1]. Uganda is estimated to have contributed 12.6 million cases and 2.7% of Africa’s 569,000 malaria-related deaths in 2023 [1]. Risks for malaria and poor malaria outcomes are greatest in young children and pregnant women.

Malaria during pregnancy can be accompanied by placental sequestration of *P. falciparum*-infected erythrocytes, leading to complications including intrauterine growth restriction, preterm delivery, and low birth weight [2]. To help control malaria in pregnancy, the World Health Organization has since 2014 recommended monthly intermittent preventive therapy with sulfadoxine-pyrimethamine (IPTp-SP) starting in the second trimester in regions with moderate to high malaria transmission [1]. IPTp-SP, adopted by 34 sub-Saharan African countries, including Uganda, was shown in studies conducted many years ago to reduce maternal anemia, antenatal and placental parasitemia, reproductive tract infections, and low birth weight pregnancies [3]. However, there is concern that, more recently, widespread resistance of *P. falciparum* to SP has compromised its preventive efficacy, especially in East and Southern Africa [4].

*P. falciparum* resistance to SP is driven by mutations in the *pfdhfr* and *pfdhps* genes, which encode the enzyme targets of pyrimethamine and sulfadoxine, respectively [5]. Multiple mutations impact on SP susceptibility in a step-wise manner. In most of East and Southern Africa, the quintuple mutant, comprising 3 PfDHFR (N51I, C59R, S108N) and 2 PfDHPS (A437G, K540E) mutations, has been associated with reduced ability of SP to clear circulating parasites and prevent new infections during pregnancy [6]. Absence of one of these 5 mutations, PfDHPS K540E, in much of West Africa appears to be associated with improved preventive efficacy of SP [4].

In the setting of the quintuple mutant haplotype, addition of the PfDHFR I164L and/or PfDHPS A581G mutations is associated with higher-level SP resistance [5, 7]. These mutations have emerged in eastern Africa, including Uganda, Tanzania, and Malawi; were selected in women receiving IPTp [8, 9]; and have been associated with reduced SP preventive efficacy [10-12]. Two meta-analyses showed that IPTp-SP preventive efficacy was greatly reduced in the setting of ≥90% prevalence of the PfDHPS K540E mutation or ≥10% prevalence of the PfDHPS A581G mutation [13, 14].

With increasing resistance to SP, the artemisinin-based combination therapy (ACT) dihydroartemisinin-piperaquine (DP) has been considered as a replacement for SP in IPTp [2]. Recent trials showed significant reductions in the incidence of malaria, parasite prevalence, and parasitemia at delivery with IPTp-DP compared to IPTp-SP [6, 15, 16]. Dihydroartemisinin (DHA) is the active metabolite of the artemisinin component of each approved ACT. Artemisinins are threatened by artemisinin partial resistance (ART-R), manifest as delayed parasite clearance after therapy and mediated by specific mutations in the PfK13 propeller domain [17]. ART-R was first identified in Southeast Asia, and has emerged and spread over the last decade in parts of eastern Africa, including Uganda [18]. Piperaquine is a bisquinoline with the longest half-life of approved ACT partner drugs and a prolonged post-treatment prophylactic effect. Mutations associated with resistance to chloroquine and amodiaquine (PfCRT K76T and PfMDR1 N86Y) have been associated with decreased piperaquine susceptibility and selected by prior treatment with DP in some studies, but these mutations are now very uncommon in most of Uganda [19]. In Southeast Asia, but not to date in Africa, novel mutations in PfCRT and duplication of plasmepsin genes have been associated with resistance to piperaquine [20]. Resistance to both artemisinins and piperaquine has been accompanied by frequent treatment failures with DP [21], but to date, DP treatment responses have remained excellent in Uganda [22].

To add to our understanding of impacts of IPTp regimens on the clearance and prevention of malarial parasitemia and selection of drug resistance mutations, we evaluated parasites collected from participants in a recent trial comparing IPTp outcomes for SP, DP, or DP+SP.

## MATERIALS AND METHODS

### Study Design

We utilized samples collected from women enrolled from December, 2020 to December, 2023 in a randomized, double-blinded controlled trial comparing monthly IPTp with SP vs. DP vs. DP+SP in Busia District, Uganda, an area of intense, perennial malaria transmission (NCT04336189). Details of the parent trial have been published elsewhere [16]. Briefly, HIV-uninfected pregnant women were enrolled at 12-20 weeks gestation, randomized to one of the study arms, and followed every 28 days through delivery for administration of study drugs and blood sample collection (routine visits). In addition, participants were encouraged to come to a dedicated study clinic open 7 days/week any time they were ill (unscheduled visits). At enrollment and during subsequent routine or unscheduled visits, participants with fever and a blood smear positive for malaria parasites were administered artemether-lumefantrine or, for complicated malaria, intravenous artesunate. Blood samples collected at routine visits or unscheduled visits with febrile illness were assessed with Giemsa-stained thick blood smears, and blood samples collected at routine visits were further evaluated by quantitative PCR (qPCR) targeting the *P. falciparum* multicopy conserved *var* gene acidic terminal sequence [23].

This study had 3 objectives. First, to evaluate the efficacy of IPTp drugs in clearing parasites and preventing new infections, we compared risks of recurrent parasitemia in participants with asymptomatic parasitemia (positive by blood smear with parasite density >100/µL by qPCR) at the time of each administration of the different IPTp regimens. Second, to assess the selection of drug resistance markers following IPTp, we evaluated associations between the number of prior doses of the different IPTp regimens received and known markers of resistance to SP and DP. Third, to assess whether markers of drug resistance were associated with failure to clear parasites, in participants in the IPTp-SP arm, we evaluated associations between the presence of mutations associated with high-grade SP resistance and the risk of recrudescent parasitemia.

### Ethics

Informed consent for storage and future use of biological specimens was obtained at the time of enrollment in the parent IPTp study. The study was approved by the Makerere University School of Biomedical Sciences Research Ethics Committee, the Makerere University School of Medicine Research Ethics Committee, the Uganda National Council for Science and Technology, and the University of California, San Francisco Committee on Human Research.

### Sequencing of *P. falciparum* Isolates

Genomic DNA was extracted from whole blood samples using the Pure Link *Pro* 96 genomic DNA purification kit (Invitrogen) following the manufacturer’s instructions, and diluted 1:4 in water. *P. falciparum* genetic polymorphisms of interest in the *pfdhfr, pfdhps, pfcrt*, pfmdr1, and *pfk13* genes were characterized using the MAD^4^HatTeR deep sequencing platform [24, 25]. Samples were genotyped at targets in the D1/R1.2 +⍰R2 primer pools encompassing common drug resistance mutations and 165 diversity loci using the experimental conditions detailed in V.4 of the MAD^4^HatTeR protocol (https://www.protocols.io/view/mad4hatter-14egn779mv5d/v4). Sequencing was performed on the Illumina NextSeq platform and generated reads were analyzed using the MAD^4^HatTeR bioinformatic pipeline version 0.2.1 (https://github.com/EPPIcenter/mad4hatter).

### Genomic Data Analysis

Genotypes were called if the allele had ≥10 reads and within-sample allele frequency (WSAF) ≥1%. Unless otherwise indicated, described mutant prevalences included mixed and pure mutant samples. For determination of *plasmepsin* gene copy number, we applied a generalized additive model to normalize read depth and estimate fold change across targets in the *plasmepsin* 2 and *plasmepsin* 3 genes using *P. falciparum* 3D7 as the single-copy reference [25]; the cut-off for increased copy number was >1.6 copies.

To distinguish recrudescent from new infections, we used diversity microhaplotype data from MAD^4^HatTeR and the Adaptive Statistical Framework for Therapeutic Efficacy and Recrudescence (Aster), a likelihood-based method that uses identity-by-descent analysis to detect persistent clones and accounts for alleles matching by chance [26]. For inclusion in the analysis, a sample was required to have >100 reads for >10% of the diversity microhaplotypes. Recrudescent infections were identified when paired samples included a persistent strain, utilizing the R package *asterTES* [26].

To compare parasite strains across multiple timepoints, we phased microhaplotype data over time using a multi-step approach. At each timepoint, we filtered loci to retain microhaplotypes with WSAF ≥0.7. Timepoints with a single microhaplotype per locus after filtering defined the initial strain pool. Each strain was then queried across all timepoints for a study participant. A strain was considered present when all its defining microhaplotypes were detected, with its frequency assigned as the minimum among these microhaplotypes. When a strain was detected, we subtracted its frequency from the corresponding microhaplotypes. If this subtraction left one microhaplotype per locus, we inferred an additional strain. Finally, all strains were re-evaluated across timepoints to determine presence and within-sample frequencies. Strain-specific parasite densities were calculated by multiplying within-sample frequencies by total parasite density.

### Statistical analysis

Data were analyzed using STATA version 17, with a two-sided p-value of 0.05 considered statistically significant. For objective 1, we assessed recurrent and genotype-confirmed recrudescent microscopic parasitemia over 28-days after each administration of IPTp drugs, including episodes with symptoms requiring antimalarial therapy. The cumulative risks of each outcome were estimated using the Kaplan-Meier product limit formula with 95% confidence intervals. Associations between IPTp regimens and treatment outcomes were assessed using a Cox proportional hazards model with adjustment for repeated measures in the same study participant and expressed as hazard ratios (HRs). Analyses were also stratified by gravidity. For objective 2, associations between the number of prior doses of IPTp and resistance mutations were assessed using generalized estimating equations to adjust for repeated measures in the same participant with a log-binomial model and robust standard errors to obtain risk ratios with 95% confidence intervals. For objective 3, we assessed associations between resistance markers and recrudescent parasitemia using the statistical methods described for objective 1.

## RESULTS

### Study samples

A total of 1405 samples fulfilled our selection criteria, of which 1377 (98%) were successfully genotyped and included in the analysis; 771 were collected at the time the first IPTp dose was administered and 606 at later timepoints (Figure 1). At initiation of IPTp the mean age of study participants with asymptomatic parasitemia was 22.3 years, with mean gestational age 17.4 weeks, and 47.5% of participants primigravidae. Mean parasite densities were 781 and 1016 parasites/µL by microscopy and qPCR, respectively (Table 1).

**Table 1.**
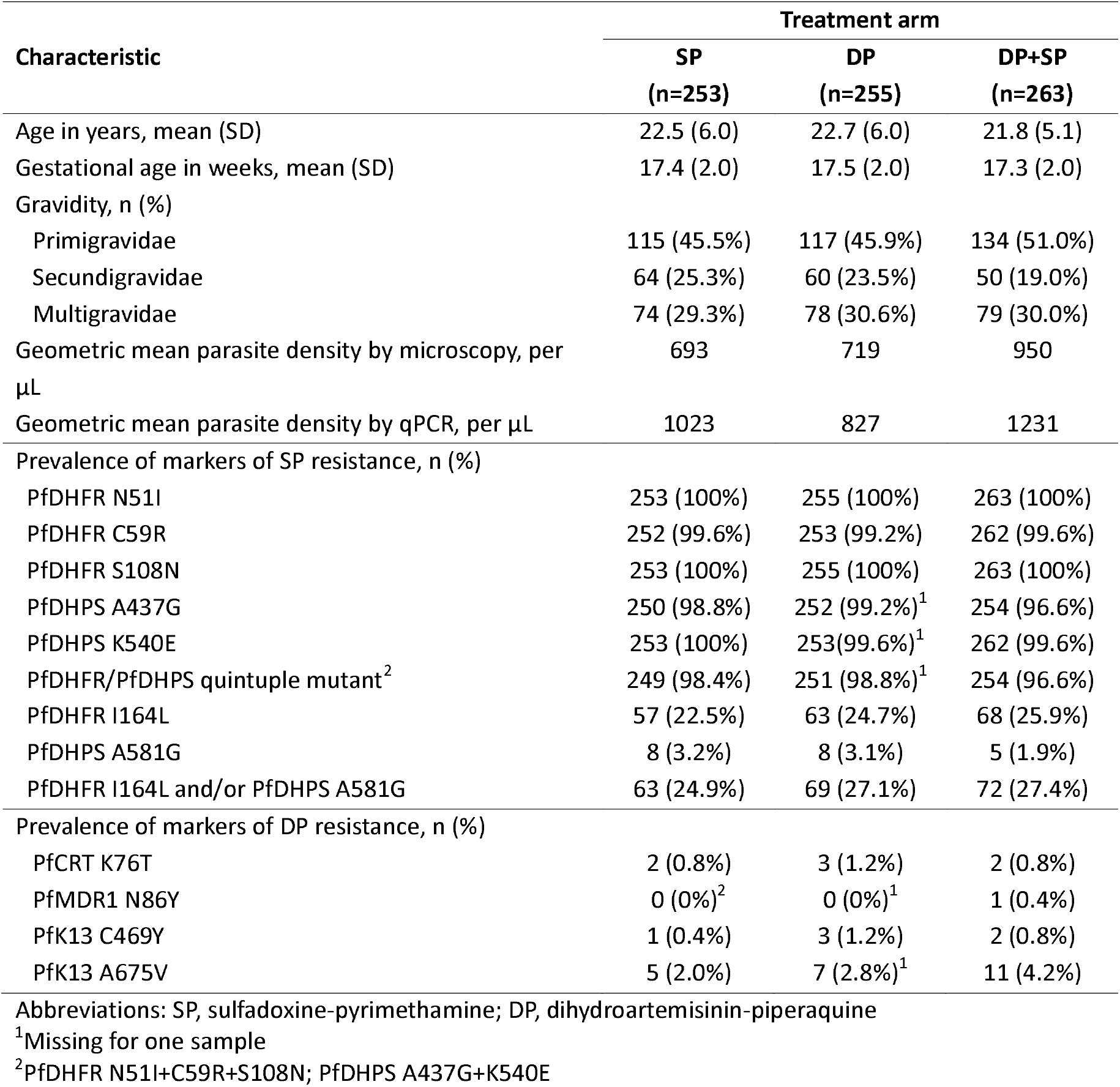
Characteristics of study participants from samples collected at initiation of IPTp.

**Figure 1.**
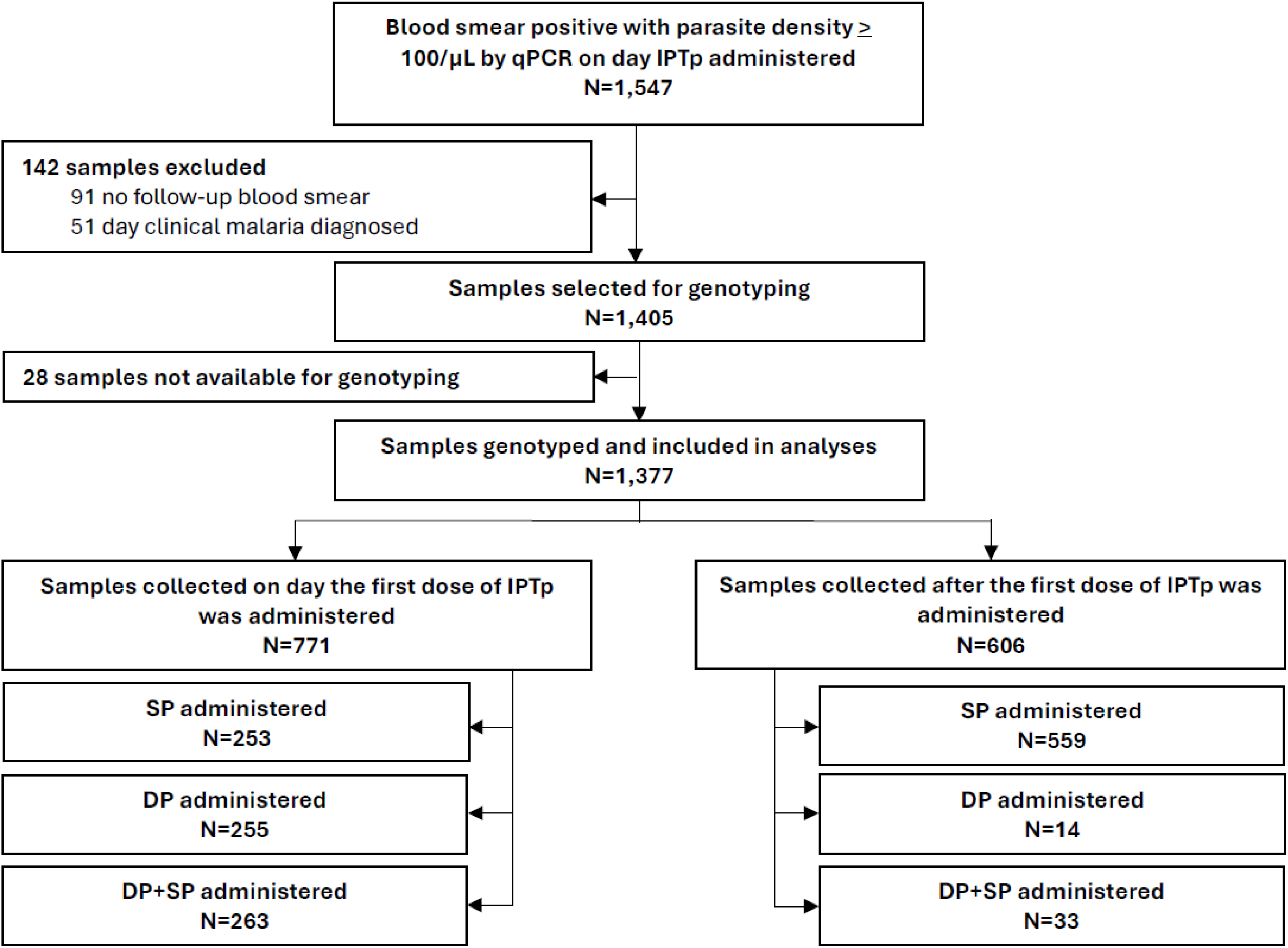
Study profile.

### Drug resistance polymorphisms in samples collected at initiation of IPTp

The combined prevalence of 5 well-characterized SP-resistance mutations (PfDHFR N51I, C59R, S108N; PfDHPS A437G, K540E), which comprise the quintuple mutant genotype, was 97.9% (Table 1). Prevalences of the I164L and A581G mutations, which are associated with higher-level SP resistance, were 24.4% and 2.7%, respectively; 26.5% of samples had at least one of these two mutations. Prevalences of the PfCRT K76T, PfMDR1 N86Y, PfK13 C469Y, and PfK13 A675V mutations, which are potentially associated with decreased susceptibility to DHA or piperaquine, were all <5% (Table 1). Considering polymorphisms associated with piperaquine resistance in Southeast Asia, the PfCRT H97Y, C101F, F145I, M343L, and G353V mutations [27, 28] and increased *plasmepsin 2/3* gene copy number [29, 30] were not seen in any samples.

### Associations between IPTp regimens and risks of recurrent parasitemia

The cumulative risks of any recurrent parasitemia (SP 57.8%, DP 4.1%, DP+SP 3.9%) and symptomatic malaria (SP 9.3%, DP 1.1%, DP+SP 0.3%) were significantly greater in the SP treatment arm (Table 2). After genotyping to distinguish new and recrudescent parasitemia, the cumulative risk of recrudescent parasitemia (persistent infection with the same *P. falciparum strain*) was also significantly greater in the SP treatment arm (SP 40.1%, DP 2.0%, DP+SP 0.8%). In some participants, especially primigravid women, the inability of IPTp-SP to clear parasitemia was striking. Of 156 primigravid women in the IPTp-SP arm included in this study, 125 (80.1%) experienced at least one additional episode of recurrent parasitemia despite monthly IPTp-SP, with 102 (65.4%) experiencing at least one episode of recrudescence (persistence of a strain following administration of IPTp-SP). In 9 subjects, recrudescence was seen in at least 4 consecutive samples collected over the course of pregnancy. One participant had persistence (without symptoms) of a baseline strain in all of 6 subsequent samples evaluated over 6 months until shortly before delivery (Figure 2). Five other subjects had 3-5 consecutive monthly recrudescent samples; others had intermittent reappearance of the same recrudescent isolate over the course of pregnancy (Figure 2).

**Table 2.** Risk of recurrent parasitemia following administration of different IPTp regimens.

| IPTp regimen | All infections |  |  | Recrudescent infections |  |  |
| --- | --- | --- | --- | --- | --- | --- |
|  | Cumulative risk of recurrent parasitemia (95% CI) | HR (95% CI) | p-value | Cumulative risk of recurrent parasitemia (95% CI) | HR (95% CI) | p-value |
| ***Any parasitemia during 28-day follow-up*** |  |  |  |  |  |  |
| SP (n=812) | 57.8% (54.3-61.3%) | Reference group |  | 40.1% (36.7-43.8%) | Reference group |  |
| DP (n=269) | 4.1% (2.2-7.4%) | 0.07 (0.03-0.15) | <0.001 | 2.0% (0.8-4.7%) | 0.05 (0.01-0.19) | <0.001 |
| DP+SP (n=296) | 3.9% (2.1-7.4%) | 0.06 (0.03-0.11) | <0.001 | 0.8% (0.2-3.1%) | 0.02 (0.005-0.08) | <0.001 |
| ***Parasitemia during 28-day follow-up with symptoms requiring antimalarial therapy*** |  |  |  |  |  |  |
| SP (n=812) | 9.3% (7.4-11.5%) | Reference group |  | 6.9% (5.3-8.9%) | Reference group |  |
| DP (n=269) | 1.1% (0.4-3.4%) | 0.12 (0.04-0.38) | <0.001 | 0.8% (0.2-3.0%) | 0.11 (0.03-0.45) | 0.002 |
| DP+SP (n=296) | 0.3% (0.1-2.4%) | 0.04 (0.01-0.26) | 0.001 | 0.3% (0.1-2.4%) | 0.05 (0.01-0.36) | 0.003 |

**Figure 2.**
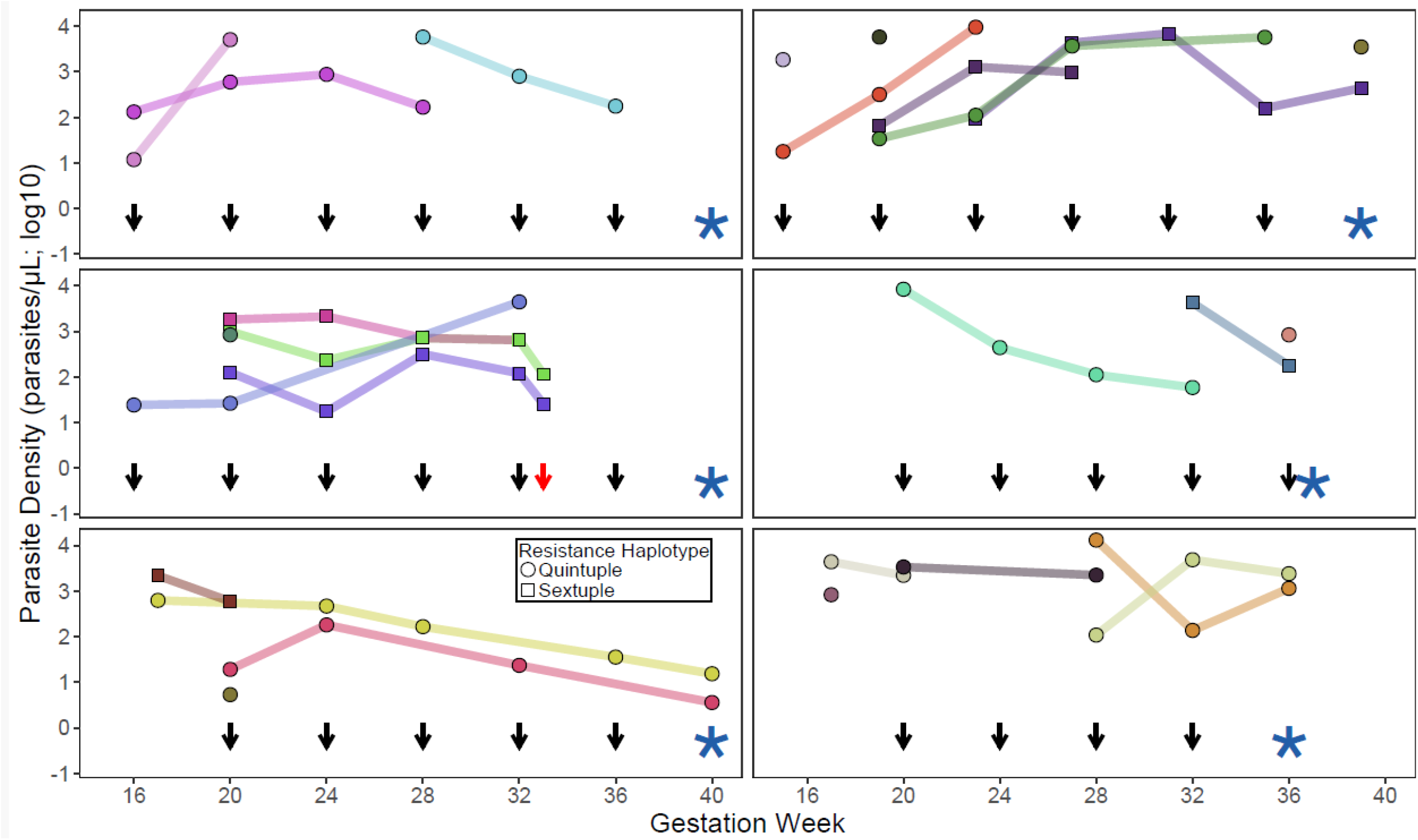
Parasitemia over the course of pregnancy in 6 selected research participants. Different isolates, as characterized with Aster, are shown in different colors; persistent isolates between timepoints (i.e. recrudescences) are indicated by lines. The shapes indicate the presence of 5 or 6 antifolate resistance mutations, as defined in the key. Parasite densities are based on qPCR quantification. Black arrows represent dates of administration of IPTp-SP; the red arrow indicates the date of administration of artemether-lumefantrine for symptomatic malaria. Stars represent dates of delivery.

Among participants receiving IPTp-SP, the risk of recurrent parasitemia during follow-up was strongly associated with gravidity. Compared to multigravidae, primigravidae had a significantly higher risk of any recurrent parasitemia (76.9% vs. 32.7%; p<0.001) and recrudescent parasitemia (54.6% vs. 24.4%; p<0.001), with a step-wise decrease in the risk of parasitemia with increasing gravidity (Supplementary Table 1, Figure 3). Similarly, primigravidae had higher risks of symptomatic malaria (Supplementary Table 1). Significant associations were not observed between gravidity and risks of parasitemia or symptomatic malaria among women receiving IPTp with DP or DP+SP.

**Figure 3.**
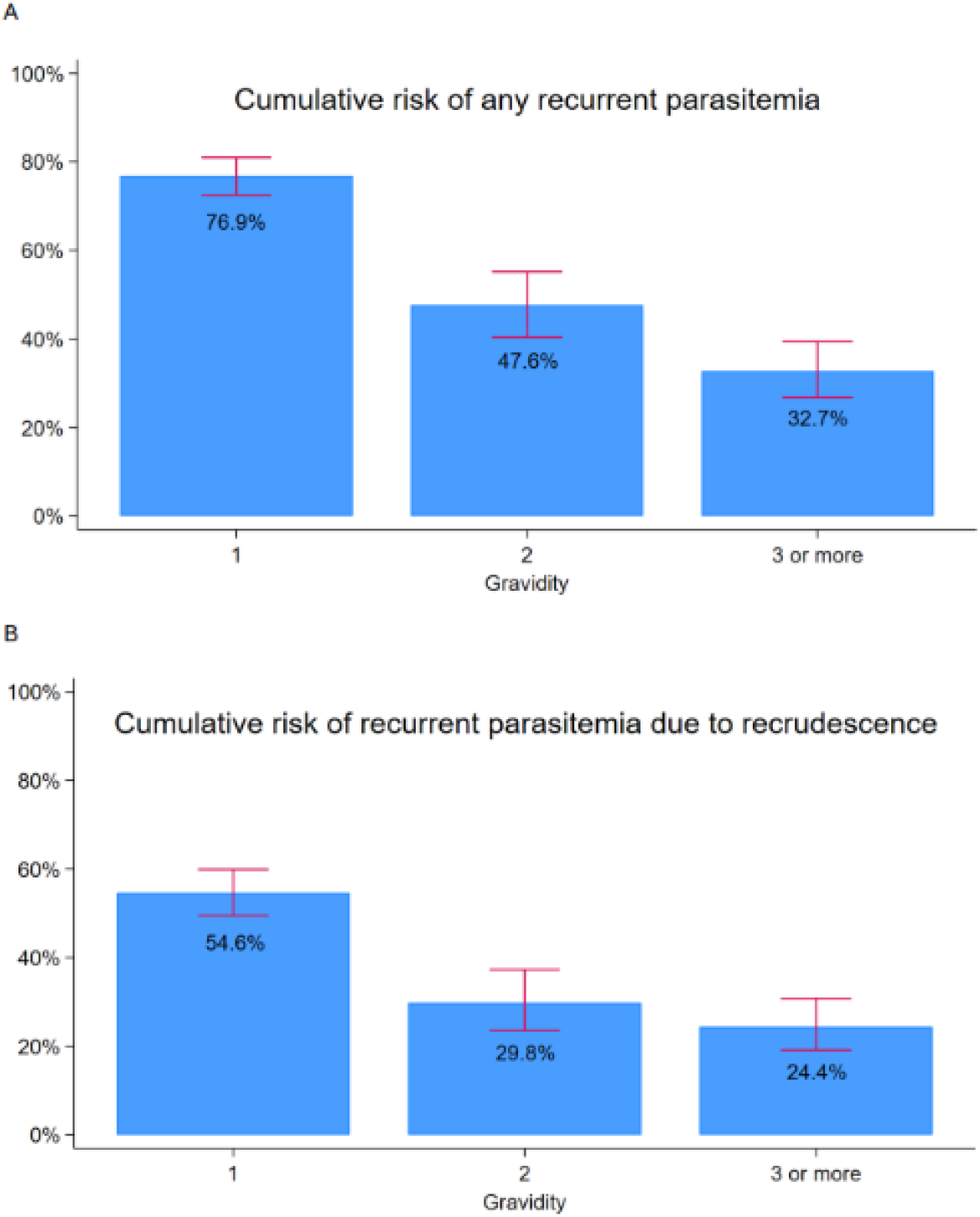
Risks of recurrent parasitemia with different gravidities. Cumulative risks of any recurrent parasitemia (A) and recurrence due to recrudescence (B) are shown. Error bars represent 95% confidence intervals.

### Selection of drug resistance markers by IPTp regimens

The prevalence of high-level SP resistance markers (PfDHFR I164L and/or PfDHPS A581G) increased from 26.5% before initiation of IPTp to 33.8% among all samples collected after initiation (relative risk [RR] 1.23, 95% CI: 1.02-1.48, p=0.03). When stratified by IPTp regimen, the prevalence of these two mutations increased from 24.9% before initiation of IPTp-SP to 35.2% after initiation (RR 1.42; 95% CI: 1.12-1.79, p=0.004), without significant changes in prevalences in the other study arms (Table 3). A dose response was seen, with increasing prevalences of the two mutations with increasing numbers of doses of IPTp-SP, but not the other study regimens (Table 3). The relative risks for these mutations were even higher when considering risks of only pure (rather than mixed and pure) mutant infections (Supplementary Table 2). Mutations in PfCRT and PfMDR1 associated with resistance to aminoquinolines and in PfK13 associated with ART-R were uncommon; there was no evidence of selection of these mutations by study regimens, although small numbers of parasite-positive samples from the DP arms of the study limited the statistical power for this analysis (Supplementary Table 3). The polymorphisms associated with piperaquine resistance in Southeast Asia noted above were not seen in any samples collected after initiation of IPTp.

**Table 3.** Selection of molecular markers associated with high-grade SP resistance^1^.

| IPTp regimen | Number of prior doses | DHFR I164L |  |  | DHPS A581G |  |  | DHFR I164L and/or DHPS A581G |  |  |
| --- | --- | --- | --- | --- | --- | --- | --- | --- | --- | --- |
|  |  | Prevalence | RR (95% CI)<sup>1</sup> | p-value | Prevalence | RR (95% CI)<sup>1</sup> | p-value | Prevalence | RR (95% CI)<sup>1</sup> | p-value |
| SP | 0 | 57/253 (22.5%) | Reference group |  | 8/253 (3.2%) | Reference group |  | 63/253 (24.9%) | Reference group |  |
|  | 1-3 | 129/416 (31.0%) | 1.37 (1.06-1.77) | 0.02 | 18/416 (4.3%) | 1.58 (0.71-3.53) | 0.27 | 138/416 (33.2%) | 1.35 (1.06-1.72) | 0.02 |
|  | 4-6 | 51/143 (35.7%) | 1.57 (1.15-2.14) | 0.004 | 14/143 (9.8%) | 3.07 (1.29-7.30) | 0.01 | 59/143 (41.3%) | 1.62 (1.23-2.14) | 0.001 |
|  | Any (1-6) | 180/559 (32.2%) | 1.41 (1.10-1.82) | 0.007 | 32/559 (5.7%) | 1.96 (0.92-4.18) | 0.08 | 197/559 (35.2%) | 1.42 (1.12-1.79) | 0.004 |
| DP | 0 | 63/255 (24.7%) | Reference group |  | 8/255 (3.1%) | Reference group |  | 69/255 (27.1%) | Reference group |  |
|  | 1-3 | 1/9 (11.1%) | 0.94 (0.49-1.78) | 0.84 | 0/9 (0%) | NA |  | 1/9 (11.1%) | 0.83 (0.42-1.63) | 0.59 |
|  | 4-6 | 1/5 (20.0%) | 0.88 (0.19-4.15) | 0.87 | 0/5 (0%) | NA |  | 1/5 (20.0%) | 0.77 (0.16-3.84) | 0.75 |
|  | Any (1-6) | 2/14 (14.3%) | 0.91 (0.48-1.71) | 0.77 | 0/14 (0%) | NA |  | 2/14 (14.3%) | 0.80 (0.42-1.55) | 0.52 |
| DP+SP | 0 | 68/263 (25.9%) | Reference group |  | 5/263 (1.9%) | Reference group |  | 72/263 (27.4%) | Reference group |  |
|  | 1-3 | 3/18 (16.7%) | 0.61 (0.20-1.87) | 0.39 | 1/18 (5.6%) | 2.94 (0.36-23.7) | 0.31 | 4/18 (22.2%) | 0.74 (0.28-1.99) | 0.55 |
|  | 4-6 | 1/15 (6.7%) | 0.30 (0.06-1.50) | 0.14 | 1/15 (6.7%) | 3.53 (0.44-28.4) | 0.24 | 2/15 (13.3%) | 0.55 (0.18-1.71) | 0.30 |
|  | Any (1-6) | 4/33 (12.1%) | 0.48 (0.19-1.20) | 0.12 | 2/33 (6.1%) | 3.21 (0.65-15.9) | 0.15 | 6/33 (18.2%) | 0.66 (0.31-1.40) | 0.28 |

<sup>1</sup>Adjusted for repeated measures in the same study participant

### Association between high-level SP resistance markers and recrudescent infections

We explored whether the PfDHFR I164L or PfDHPS A581G mutations were associated with risks of recrudescent infections after IPTp-SP. Infection with parasites containing either the PfDHFR I164L or PfDHPS A581G mutation was associated with modest, but not statistically significant increases in the subsequent risks of recrudescent parasitemia or symptomatic malaria (Table 4).

**Table 4.** Associations between molecular markers associated with high-grade SP resistance and risk of recrudescent parasitemia following IPTp-SP.

| Genotype |  | Any parasitemia due to recrudescence during 28-day follow-up |  |  | Parasitemia due to recrudescence with symptoms requiring antimalarial therapy during 28-day follow-up |  |  |
| --- | --- | --- | --- | --- | --- | --- | --- |
|  |  | Cumulative risk of parasitemia (95% CI) | HR (95% CI) | p-value | Cumulative risk of parasitemia (95% CI) | HR (95% CI) | p-value |
| DHFR I164L | Absent (n=575) | 38.5% (34.4-42.9%) | Reference group |  | 6.6% (4.8-9.0%) | Reference group |  |
|  | Present (n=237) | 43.9% (37.6-50.8%) | 1.17 (0.94-1.46) | 0.16 | 7.5% (4.8-11.9%) | 1.13 (0.65-1.95) | 0.68 |
| DHPS A581G | Absent (n=772) | 39.6% (36.1-43.4%) | Reference group |  | 6.6% (5.0-8.6%) | Reference group |  |
|  | Present (n=40) | 50.4% (35.3-67.6%) | 1.37 (0.86-2.17) | 0.19 | 12.5% (5.4-27.5%) | 2.08 (0.87-4.98) | 0.10 |
| DHFR I164L and/or DHPS A581G | Absent (n=552) | 38.7% (34.5-43.2%) | Reference group |  | 6.5% (4.7-8.9%) | Reference group |  |
|  | Present (n=260) | 43.1% (37.0-49.6%) | 1.14 (0.92-1.42) | 0.22 | 7.6% (4.9-11.7%) | 1.17 (0.68-2.00) | 0.58 |

## DISCUSSION

SP is widely used for malaria chemoprevention, but its value is seriously challenged by resistance to both components of the regimen. DP is a promising alternative for chemoprevention, but resistance to its components is also a potential concern. We utilized samples from a recent randomized trial [16] to compare the abilities of IPTp with SP, DP, and DP+SP to clear parasitemia, prevent new infections, and select for drug resistance markers. In the setting of 5 or more known SP resistance-mediating mutations in PfDHFR and PfDHPS, IPTp-SP performed poorly in clearing infections or preventing new infections, and it selected for additional mutations linked to high-level resistance. Thus, the antimalarial benefits of IPTp-SP appear to be minimal.

Resistance to SP is mediated by mutations in the target PfDHFR and PfDHPS enzymes, with, in Uganda, 5 mutations associated with decreased antimalarial activity (commonly referred to as the quintuple mutant) nearly fixed, and increasing prevalence also of the PfDHFR I164L and PfDHPS A581G mutations, which mediate high-level resistance [19]. In our randomized trial, at initiation of IPTp 38% of study participants had parasites detected by microscopy, 98% of these had the quintuple mutant haplotype, and 27% had in addition the I164L and/or A581G mutations. In this setting the preventive efficacy of IPTp-SP was poor. Considering participants who were parasitemic before the initiation of IPTp, risks of recurrent parasitemia and symptomatic malaria were much greater in the IPTp-SP arm compared to the DP arms. A majority of the recurrent episodes were recrudescences, i.e. the same strain caused multiple episodes of parasitemia despite IPTp-SP, indicating poor efficacy for the elimination of circulating parasites. Among primigravid women, who lack gravidity-dependent immunity and are at the greatest risk of malaria, the risk of any recurrent parasitemia was 77% and the risk of recrudescent parasitemia was 55% following administration of IPTp-SP. Thus, in Uganda, where circulating parasites typically have moderate to high-level SP resistance, the utility of IPTp-SP for malaria chemoprevention is questionable, especially in primigravidae.

Our results for SP were consistent with those seen in other studies. IPTp-SP has not been studied against a placebo since the early 2000s (before the widespread emergence of SP resistance), but its antimalarial preventive efficacy compared to IPTp-DP was poor in multiple recent studies across Africa [15, 31-35]. Compared to a prior study conducted in 2016-17 in the same region as our new study [15], baseline prevalences of the quintuple mutant haplotype and the PfDHPS A581G mutation were similar, but prevalence of the PfDHFR I164L mutation was much higher (increased from 4.0% to 24.4%), consistent with increasing spread of this mutation across Uganda over time [19]. Thus, the already poor preventive efficacy of SP is likely decreasing with spread of higher-level resistance. Studies of IPT-SP in infants [36] and schoolchildren [36] in Uganda have also shown limited efficacy. Indeed, in a randomized trial of infants 0.5-2 years of age conducted from 2010-13 in the same area of Uganda as this study, chemoprevention with monthly SP provided no protection against parasitemia or symptomatic malaria compared to no chemoprevention [36]. Our study adds insight into the ability of IPTp-SP to clear and prevent *P. falciparum* infections. In fact, IPT-SP performed poorly, with most patients in the SP arm, but <5% in DP-containing arms, remaining parasitemic in the month following a dose of IPTp.

It was of interest also to explore potential selection of resistance by IPT-DP. Partial resistance to DHA is mediated principally by known mutations in the PfK13 protein [18], and resistance to piperaquine is mediated in Southeast Asia by amplification of plasmepsin genes and novel mutations in PfCRT not previously seen in Africa [20]. In our trial, recurrent infections were uncommon in the IPTp-DP and IPT-DP+SP arms, limiting the power for consideration of resistance selection. But, in the observed recurrent infections, the prevalence of ART-R-mediating PfK13 mutations remained low and was not increased after receipt of IPTp-DP or IPTp-DP+SP, and polymorphisms associated with piperaquine resistance in Southeast Asia were not seen.

It was also of interest to determine if presence of the PfDHFR I164L or PfDHPS A581G mutations, whose prevalence is increasing in Uganda, was associated with increased risk of recrudescence after receiving IPTp-SP. Risks of recrudescence were higher for infections containing the PfDHFR I164L or PfDHPS A581G mutations, but these differences did not reach statistical significance, perhaps due to limited statistical power or the fact that the presence of the quintuple mutation is sufficient to confer a high risk of failure. Taken together, our results suggest that, in the presence of the quintuple mutant phenotype the preventive efficacy of IPTp-SP is poor, but that additional mutations that further decrease SP activity have limited additional impact on the preventive efficacy of IPTp-SP.

It is well-established that risks of malaria during pregnancy and poor birth outcomes are much greater in primigravidae than in those with prior pregnancies, and in our parent study the burden of malaria and risks of adverse birth outcomes were greatest in primigravidae. Our analysis adds the understanding that, after receipt of IPTp-SP, risks of recurrent parasitemia, recurrent symptomatic malaria, and recrudescent infection were all much greater in primigravidae. These results amplify the message that measures to control malaria during pregnancy are most relevant in primigravidae. Indeed, due to the markedly better antimalarial preventive efficacy of DP compared to SP, a switch to IPTp-DP specifically in primigravidae might be considered for areas with moderate to high malaria transmission intensity.

Our study had some limitations. First, due to ethical concerns regarding the inclusion of a placebo arm, results for IPTp-SP could only be compared to those with IPTp including DP. Thus, the absolute preventive efficacy of IPTp-SP could not be determined. Second, our study was limited to known mediators of resistance to SP and DP. Outcomes may have been impacted by, and study regimens may have selected for other resistance mediators not considered in this analysis. Third, the study was conducted in an area with very high malaria transmission intensity and high prevalence of mutations mediating moderate-to-high level resistance to the components of SP; generalizability to regions with lower transmission intensity and lower-level SP resistance is uncertain.

In summary, IPTp-SP offered disappointing antimalarial efficacy, and much poorer efficacy compared to IPTp-DP, in clearing parasitemia and preventing new infections in a region with a high prevalence of resistance of *P. falciparum* to SP. Malaria risks and impacts of SP resistance were greatest in primigravidae. These results suggest that, although IPTp-DP has not shown superior overall efficacy in preventing poor birth outcomes compared to IPTp-SP, a strategy using IPTp-DP in primigravidae in areas of high transmission intensity and SP resistance may have merit.

## Data Availability

All data produced in the present study are available upon reasonable request to the author

## Acknowledgments

We thank study participants, Infectious Diseases Research Collaboration staff based at the Tororo and Busia sites, the staff of Masafu General Hospital, and the Tororo-Busia community advisory board for participation in and support of this study. We specifically thank Harriet Adrama, Bakar Odongo, and Peter Olwoch for coordination of laboratory activities in Tororo, Jackie Nakasaanya for assistance with laboratory work in Tororo, and Emmanuel Arinaitwe for helpful advice. This work was supported by grant AI141308 from the National Institutes of Health to GD and PJR. JK was supported by a Fogarty International Center/National Institutes of Health training grant (TW010526).

## Figure Legends

**Supplementary Table 1.** Risk of recurrent parasitemia following administration of different IPTp regimens stratified by gravidity.

| IPTp regimen | Gravidity<sup>1</sup> | All infections |  |  | Recrudescent infections |  |  |
| --- | --- | --- | --- | --- | --- | --- | --- |
|  |  | Cumulative risk of recurrent parasitemia (95% CI) | HR (95% CI) | p-value | Cumulative risk of recurrent parasitemia (95% CI) | HR (95% CI) | p-value |
| ***Any parasitemia during 28-day follow-up*** |  |  |  |  |  |  |  |
| SP | Multigravida (n=227) | 32.7% (26.8-39.4%) | Reference group |  | 24.4% (19.1-30.8%) | Reference group |  |
|  | Secundigravida (n=195) | 47.6% (40.4-55.2%) | 1.48 (1.10-2.00) | <0.001 | 29.8% (23.5-37.3%) | 1.21 (0.82-1.80) | 0.33 |
|  | Primigravida (n=390) | 76.9% (72.5-81.0%) | 2.68 (2.10-3.42) | <0.001 | 54.6% (49.5-59.9%) | 2.43 (1.79-3.29) | <0.001 |
| DP | Multigravida (n=79) | 2.8% (0.7-10.8%) | Reference group |  | 1.3% (0.2-8.7%) | Reference group |  |
|  | Secundigravida (n=69) | 9.3% (4.3-20.0%) | 3.37 (0.58-19.5) | 0.17 | 4.7% (1.5-14.0%) | 3.41 (0.23-51.6) | 0.38 |
|  | Primigravida (n=121) | 1.8% (0.5-7.2%) | 0.63 (0.09-4.37) | 0.64 | 0.8% (0.1-5.7%) | 0.64 (0.004-10.3) | 0.75 |
| DP+SP | Multigravida (n=88) | 1.1% (0.2-7.8%) | Reference group |  | 1.1% (0.2-7.8%) | Reference group |  |
|  | Secundigravida (n=60) | 8.9% (3.4-22.0%) | 5.87 (0.69-49.7) | 0.10 | 0% | NA |  |
|  | Primigravida (n=148) | 3.6% (1.4-9.2%) | 2.36 (0.27-20.4) | 0.41 | 0.9% (0.1-6.2%) | 0.59 (0.04-9.14) | 0.71 |
| ***Parasitemia during 28-day follow-up with symptoms requiring antimalarial therapy*** |  |  |  |  |  |  |  |
| SP | Multigravida (n=227) | 4.1% (2.2-7.8%) | Reference group |  | 3.2% (1.5-6.6%) | Reference group |  |
|  | Secundigravida (n=195) | 7.5% (4.5-12.4%) | 1.82 (0.81-4.12) | 0.15 | 4.8% (2.5-9.1%) | 1.50 (0.57-4.00) | 0.41 |
|  | Primigravida (n=390) | 13.1% (10.1-17.0%) | 3.39 (1.66-6.92) | 0.001 | 10.0% (7.4-13.5%) | 3.28 (1.45-7.44) | 0.004 |
| DP | Multigravida (n=79) | 1.3% (0.2-8.7%) | Reference group |  | 1.3% (0.2-8.7%) | Reference group |  |
|  | Secundigravida (n=69) | 2.9% (0.7-11.2%) | 2.33 (0.22-24.6) | 0.48 | 1.5% (0.2-9.8%) | 1.16 (0.08-17.6) | 0.91 |
|  | Primigravida (n=121) | 0% | NA |  | 0% | NA |  |
| DP+SP | Multigravida (n=88) | 1.1% (0.2-7.8%) | Reference group |  | 1.1% (0.2-7.8%) | Reference group |  |
|  | Secundigravida (n=60) | 0% | NA |  | 0% | NA |  |
|  | Primigravida (n=148) | 0% | NA |  | 0% | NA |  |

<sup>1</sup>Multigravidae include secundigravidae and those with >2 pregnancies.

**Supplementary Table 2.** Selection of molecular markers associated with high-grade SP resistance only including pure mutant samples.

| IPTp regimen | Number of prior doses | DHFR I164L |  |  | DHPS A581G |  |  | DHFR I164L and/or DHPS A581G |  |  |
| --- | --- | --- | --- | --- | --- | --- | --- | --- | --- | --- |
|  |  | Prevalence | RR (95% CI)* | p-value | Prevalence | RR (95% CI)* | p-value | Prevalence | RR (95% CI)* | p-value |
| SP | 0 | 7/253 (2.7%) | Reference group |  | 2/253 (0.8%) | Reference group |  | 9/253 (3.6%) | Reference group |  |
|  | 1-3 | 41/416 (9.9%) | 3.52 (1.67-7.41) | 0.001 | 4/416 (1.0%) | 1.26 (0.27-5.79) | 0.77 | 43/416 (10.3%) | 2.96 (1.54-5.68) | 0.001 |
|  | 4-6 | 26/143 (18.2%) | 6.32 (2.98-13.4) | <0.001 | 6/143 (4.2%) | 5.19 (1.18-22.9) | 0.03 | 31/143 (21.7%) | 5.86 (3.04-11.3) | <0.001 |
|  | Any (1-6) | 67/559 (12.0%) | 4.17 (2.04-8.51) | <0.001 | 10/559 (1.8%) | 2.29 (0.57-9.24) | 0.24 | 74/559 (13.2%) | 3.65 (1.96-6.78) | <0.001 |

**Supplementary Table 3.** Selection of molecular markers associated with high-grade DP resistance^1^.

| IPTp regimen | Number of prior doses | PfCRT K76T |  |  | PfMDR-1 N86Y |  |  | PfK13 C469Y or A675V |  |  |
| --- | --- | --- | --- | --- | --- | --- | --- | --- | --- | --- |
|  |  | Prevalence | RR (95% CI)* | P-value | Prevalence | RR (95% CI)* | P-value | Prevalence | RR (95% CI)* | p-value |
| SP | 0 | 2/253 (0.8%) | Reference group |  | 0/252 (0%) | Reference group |  | 6/253 (2.4%) | Reference group |  |
|  | 1-3 | 1/416 (0.2%) | 0.18 (0.03-1.18) | 0.07 | 0/416 (0%) | NA |  | 16/416 (3.9%) | 1.53 (0.65-3.61) | 0.33 |
|  | 4-6 | 1/143 (0.7%) | 1.43 (0.17-12.1) | 0.74 | 0/143 (0%) | NA |  | 4/143 (2.8%) | 1.09 (0.31-3.87) | 0.89 |
|  | Any (1-6) | 2/559 (0.4%) | 0.42 (0.07-2.73) | 0.37 | 0/559(0%) | NA |  | 20/559 (3.6%) | 1.42 (0.60-3.36) | 0.42 |
| DP | 0 | 3/255 (1.2%) | Reference group |  | 0/254 (0%) | Reference group |  | 10/254 (3.9%) | Reference group |  |
|  | 1-3 | 1/9 (11.1%) | 13.3 (2.55-69.7) | 0.002 | 0/9 (0%) | NA |  | 0/9 (0%) | NA |  |
|  | 4-6 | 0/5 (0%) | NA |  | 0/5 (0%) | NA |  | 0/5 (0%) | NA |  |
|  | Any (1-6) | 1/14 (7.1%) | 7.28 (1.01-52.7) | 0.05 | 0/14 (0%) | NA |  | 0/14 (0%) | NA |  |
| DP+SP | 0 | 2/263 (0.8%) | Reference group |  | 1/263 (0.4%) | Reference group |  | 12/263 (4.6%) | Reference group |  |
|  | 1-3 | 0/18 (0%) | NA |  | 0/18 (0%) | NA |  | 2/18 (11.1%) | 2.48 (0.62-9.84) | 0.20 |
|  | 4-6 | 0/15 (0%) | NA |  | 0/15 (0%) | NA |  | 1/15 (6.7%) | 1.52 (0.23-9.81) | 0.66 |
|  | Any (1-6) | 0/33 (0%) | NA |  | 0/33 (0%) | NA |  | 3/33 (9.1%) | 2.06 (0.64-6.65) | 0.23 |

<sup>1</sup>Adjusted for repeated measures in the same study participant

